# ‘We can not just keep it in our palm’: A policy analysis of the integration of the case management of Neglected Tropical Diseases into the health system of Liberia

**DOI:** 10.1101/2025.02.06.25321795

**Authors:** Anna Wickenden, Laura Dean, Sassy Molyneux, Tiawanlyn G. Godwin-Akpan, Karsor K. Kollie, Maneesh Phillip, Zeela F Zaizay, Emerson Rogers, Emmy van der Grinten, Nana-Kwadwo Biritwum, Sally Theobald

## Abstract

This study investigates the development and translation process of a novel policy to integrate the case management of Neglected Tropical Diseases (NTDs) in Liberia’s health system. The policy responded to inequitable access and resource fragmentation challenges in NTD care, as highlighted in Liberia’s 2016 national strategic plan for the integrated Case Management of NTDs (1). This study explores Liberia’s efforts to integrate NTD case management into crucial health system components from the perspectives of multiple stakeholders engaged in the policy development and translation process.

A qualitative case study method was employed. The study examines stakeholder experiences and perspectives and utilises multiple analytical frameworks, including the Policy Analysis Triangle, the Power Cube and Network Analysis. Data triangulation led to the development of a conceptual framework that identifies key factors in effective health policy development and implementation and has five critical domains: people, place, process, politics, and power. These five domains are interconnected, dynamic and essential for translating complex health policies into practice.

The findings emphasise the need for health policies to embrace the complexities of integrating disease control programmes, calling for a shift from clinical-centric to holistic, multi-dimensional and multi-stakeholder approaches and policies. These insights contribute to global health policy-development evidence, underscoring the importance of contextually relevant and inclusive approaches to address health inequities and strengthen system sustainability.

**Author Summary:** In this paper, we explore the experiences and reflections of key stakeholders involved in the policy development and implementation of an integrated approach to the case management of NTDs in Liberia. The integrated approach was developed and articulated in the 2016 national strategic plan to address the inequity in access to NTD care through the health system in Liberia and the fragmentation and sustainability of human, technical and financial resources in providing NTD care in Liberia. We conducted a policy analysis using a qualitative case study of the NTD programme in Liberia. We examine the people, place, process, politics and power dynamics that enabled Liberia to be one of the few countries in the world to integrate the NTD care within critical health system building blocks. We triangulate our data sets and analyse the data, developing a conceptual framework that has five critical domains adapted from the Policy Analysis Triangle (1). These five domains are dynamic and contribute to successfully developing and translating complex health policies into practice. The results of the paper have broader implications:

**Embracing Complexity in Policy-Development:** Policies in low-resource settings must recognise and adapt to the complexities of integrating disease control programmes into health systems.
**Shift from biomedically focused to holistic approaches:** There is a crucial need to transition from clinically focused policies to those that encompass holistic, multi-dimensional health strategies that consider the dynamic interactions of multiple factors beyond the framework of one discipline.
**Adopting Multi-Stakeholder Strategies in Policy Development and Translation:** Effective global health policy-development requires the active involvement of a diverse range of stakeholders beyond healthcare professionals and policymakers.

## Introduction

Neglected Tropical Diseases (NTDs) represent a group of illnesses that are characterised by multi-faceted neglect. The populations NTDs predominantly affect are amongst the most overlooked globally in health care access, are often considered hard-to-reach, and experience multiple intersecting health and socio-economic barriers; investment in prevention and care programmes for these diseases is disproportionately small, relative to the number of people affected; funding and research attention for these diseases is minimal compared to other global health challenges (2, 3). Many discussions and studies related to global health policy have historically overlooked NTDs, it is not uncommon to encounter significant policy documents such as national health policies, supply chain policies, and health promotion strategic plans, amongst others, about populations in NTD endemic areas that fail to address access to and provision of NTD care through the health system (3–5).

Implementation models for NTDs mainly focus on either disease prevention through Mass Drug Administration (MDA) or NTD case management or care. NTD care, (also referred to as Morbidity Management and Disability Prevention (MMDP), Disease Management, Disability and Inclusion (DMDI), Disability Prevention and Medical Rehabilitation (DPMR) and Case Management (CM) encompasses the continuum of care, including case finding, diagnosis, treatment and rehabilitation, and can also include multiple other elements of intervention for people affected by NTDs. (6) The term case management is widely used in the Liberian context, but it should be acknowledged that it can be considered problematic, a shared document on terminology from the STOP TB Partnership, “Words Matter” (7), recommends avoiding the use of the term ‘case’ for a person.

An effective policy development and implementation process for integrating the care of people living with a neglected tropical disease (NTD) into the health system is crucial for achieving global health goals related to NTDs, universal health coverage (8, 9) and the World Health Assembly resolution on people-centred integrated health services (10). However, minimal evidence exists on how to do this effectively (2–4). Liberia’s experience in this area provides critical insights that can significantly contribute to bridging the evidence gap in this domain. It can inform a framework applicable to policy planning and translation in various contexts and on different health topics where integration is seen as a route to universal health coverage.

This policy analysis of the integrated strategy for the case management of NTDs in Liberia examines the interaction between interests, institutions and ideas in the policy development and implementation process (11), incorporating global, regional, national and county analyses (12). The policy analysis also examines the interlinkages with other overarching health systems policies and contextual changes that shaped the feasibility of the integrated plan through time.

Since the early 1990s, literature has emerged that explores the policy process and unpacks the critical aspects that should be considered in any policy analysis (14–18). There is a growing appreciation of the complexity of policy-development processes, with tools emerging for analysing and understanding how policy is developed (19, 20), the critical role of power (21–26), and how policy can be translated into practice (27, 28). Within the existing literature, authors refer to the need for more significant and in-depth policy analysis in areas of global health policy which have been neglected, encouraging highly participatory multi-stakeholder approaches and engagement in policy analysis (13–16). The proposed policy analysis will uniquely contribute to this field of work and will include an analysis and approach focusing on multiple policy process dimensions (16, 17).

Several frameworks for policy analysis have been considered for this study, including the ‘Policy Triangle Framework’ (13), the “Stages Heuristic,’ (18, 19), and “Network Analysis.” (19) The Policy Triangle Framework is the most appropriate for this research as it recognises that policy processes are not linear and considers the content of policy, actors, context and processes (13), however elements of multiple frameworks are used to complement the policy triangle.

### Neglected Tropical Disease (NTD) case management model in Liberia

While the definition of integration in the context of health systems, public health, and NTDs is contested (2, 20–23), there is a shared commonality across definitions that relates to the process of combining various elements—whether tools, diseases, activities, indicators, or processes. An integration process is inherently complex. Success in this context lies in navigating this complexity to achieve a broader goal. In 2016, Liberia took the decisive step of moving away from parallel or vertical approaches to managing NTDs that were implemented with limited integration with the routine health system mechanisms or governance, instead embarking on a policy revision and development process that consolidated treatment and care for individuals living with NTDs under one Programme (the NTD Programme) and guided by one integrated policy (16).

There is limited knowledge or empirical evidence of factors contributing to developing an integrated policy in complex health systems that can traverse the gap between policy development and implementation, especially in Fragile and Conflict Affected States (FCAS). This paper contributes to increasing knowledge in this area.

The integration of case management in Liberia and associated policy processes is also an essential example of changing policy discourse in a neglected area that warrants further exploration to identify the lessons that could be learnt for policy-development in Liberia, in other contexts and on other health topics.

## Methods

We employed a qualitative case study design to investigate the policy development and translation process through the experiences of 20 purposively selected stakeholders, each with a significant role in either the development or implementation of Liberia’s integrated NTD policy or in the decisions and development that precipitated the policy shift. This case study method enabled an intensive, in-depth investigation into policy development, policy implementation and health system adaptation in its real-life context through Liberia’s integrated case management model (11, 24). This case study is an example of a national government of a fragile state seeking to integrate vertical health services into the broader health system to improve access to healthcare (24).

### Sampling

This study is concerned with exploring the experiences and reflections of key stakeholders involved in the policy development and implementation of an integrated approach to the case management of NTDs in Liberia, recognising that there are multiple views of reality influenced by social context and the environment of key informants, as such the naturalist paradigm informs this qualitative study. This study follows the principles of purposive sampling guided by context and research questions. Key informants were purposively selected through stakeholder analysis based on tacit knowledge, participant and interviewee recommendations. Key informants were drawn from multiple levels of the health system recognising the reach of the policy and the diversity of experience of policy development and implementation: the national level, the county level, and the global level, according to their experience linked to their job role and knowledge of the NTD programme, NTD policy, and broader relevant health and social issues in Liberia.

For a detailed account of the participant characteristics, please refer to Table 1 below.

**Table 1.** Participant Characteristics.

| Participant Characteristics |  |  |  |  |
| --- | --- | --- | --- | --- |
| Key Informants | Variables | Categories | Count | Per cent |
| Key Informant Interviews<br>(n=20) | Sex | Female | 3 | 15% |
|  |  | Male | 17 | 85% |
|  | Employment | Government | 13 | 65% |
|  |  | Non-Government | 7 | 35% |
|  | Health system level of primary focus | International | 6 | 30% |
|  |  | National | 8 | 40% |
|  |  | County | 6 | 30% |
|  | Age | <45 | 16 | 80% |
|  |  | >/ 45 | 4 | 20% |

Participant selection was informed by a participatory stakeholder analysis workshop, which was part of the broader REDRESS research project. REDRESS is a cross-disciplinary research project, led by the Liverpool School of Tropical Medicine which aims to help improve the care of people affected by severe stigmatising skin diseases (SSSDs – a term specifically used in the research call) in Liberia, namely, leprosy, onchocerciasis, lymphatic filarisis, Buruli ulcer, and yaws, which are included in the integrated NTD case management policy. The adapted process for the stakeholder analysis with REDRESS co-investigators engaging with the integrated case management policy is outlined below. (Figure 1)

**Fig 1.**
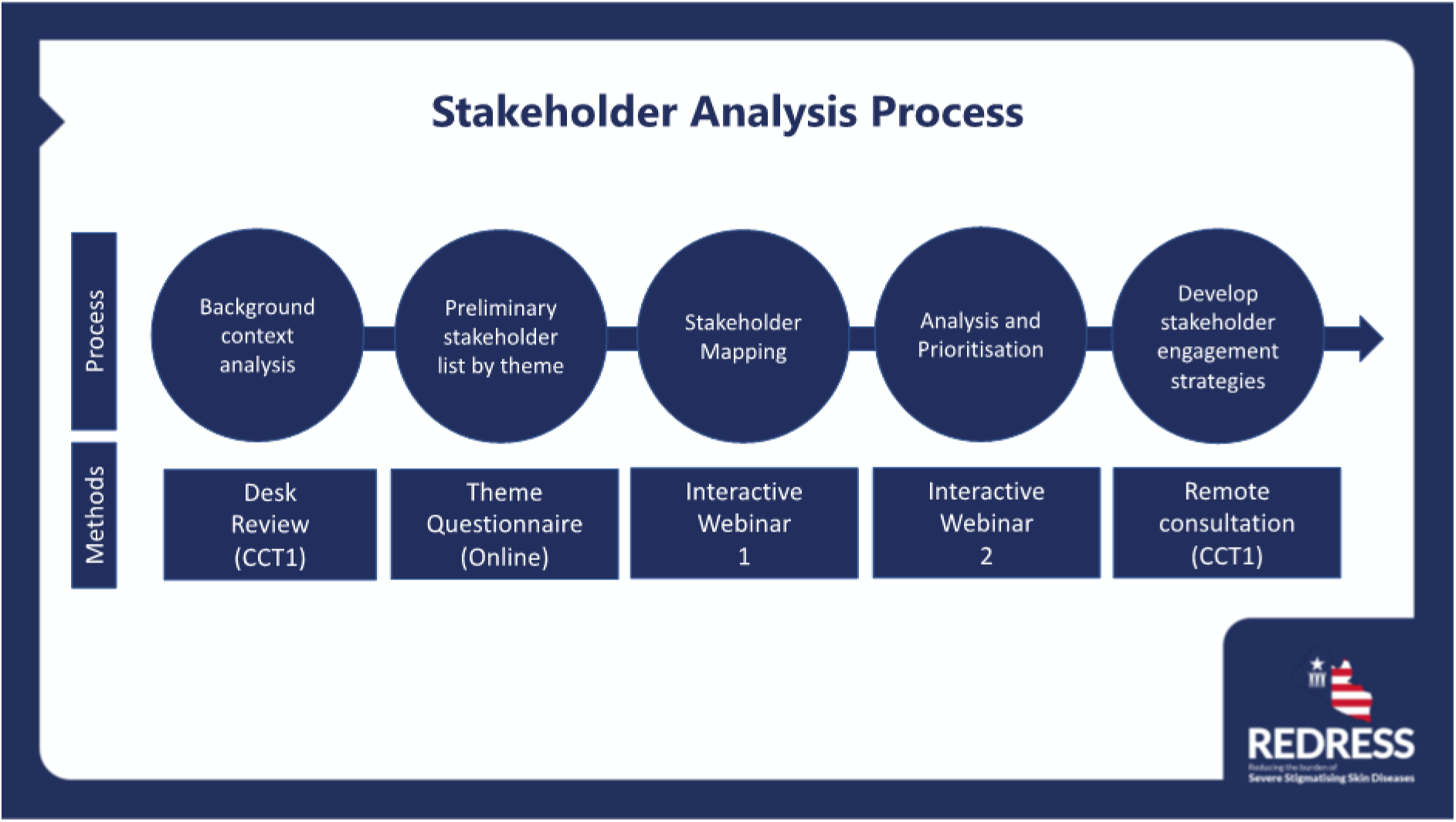
Adapted Stakeholder Analysis Process.

### Data collection

Data was collected through key informant interviews. These took place from March to December 2023, including interviews with several co-authors, all interviews were conducted by A.W. Interviews were conducted in English, and the length varied from 30 minutes to 2 hours. The interview format was exploratory, using a topic guide for a structure that explored multiple facets of the policy development process, personal reflections and pressures on its implementation. However, questions were tailored by the interviewer based on each interviewee’s context and reactions, fostering a richer narrative and reflexivity in their responses. The interviews were held in Liberia and internationally with additional interviews conducted online via video conferencing platforms (Microsoft Teams).

### Data Analysis

Interviews were transcribed verbatim, and the transcripts were stored and analysed using NVivo12 software. We applied a reflexive thematic analysis, following the six-phase method described by Braun and Clarke (25). Initially, we familiarised ourselves with the data, reviewing transcripts and creating mind maps to highlight emerging key points. The second phase involved open coding, systematically reviewing the whole dataset to identify segments relevant to our study’s focus. In the third phase, we formulated initial themes, recognising patterns and shared meanings across the dataset. We grouped clusters of codes that seemed to converge around a central idea or concept that could significantly contribute to fulfilling the aim of the study. The fifth phase entailed refining, defining, and naming our themes, ensuring each was demarcated and centred on a robust core concept. The final phase—phase six—included composing the study’s write-up, considering familiarisation diagrams and reflexive journaling that commenced at the analysis’s outset.

To conduct the analysis, the policy triangle framework (13) informed the analytical process alongside elements of the Power Cube (22, 23) and Network Analysis (26–28). Figure 2 shows the process of data collection, analysis and results.

**Fig 2.**
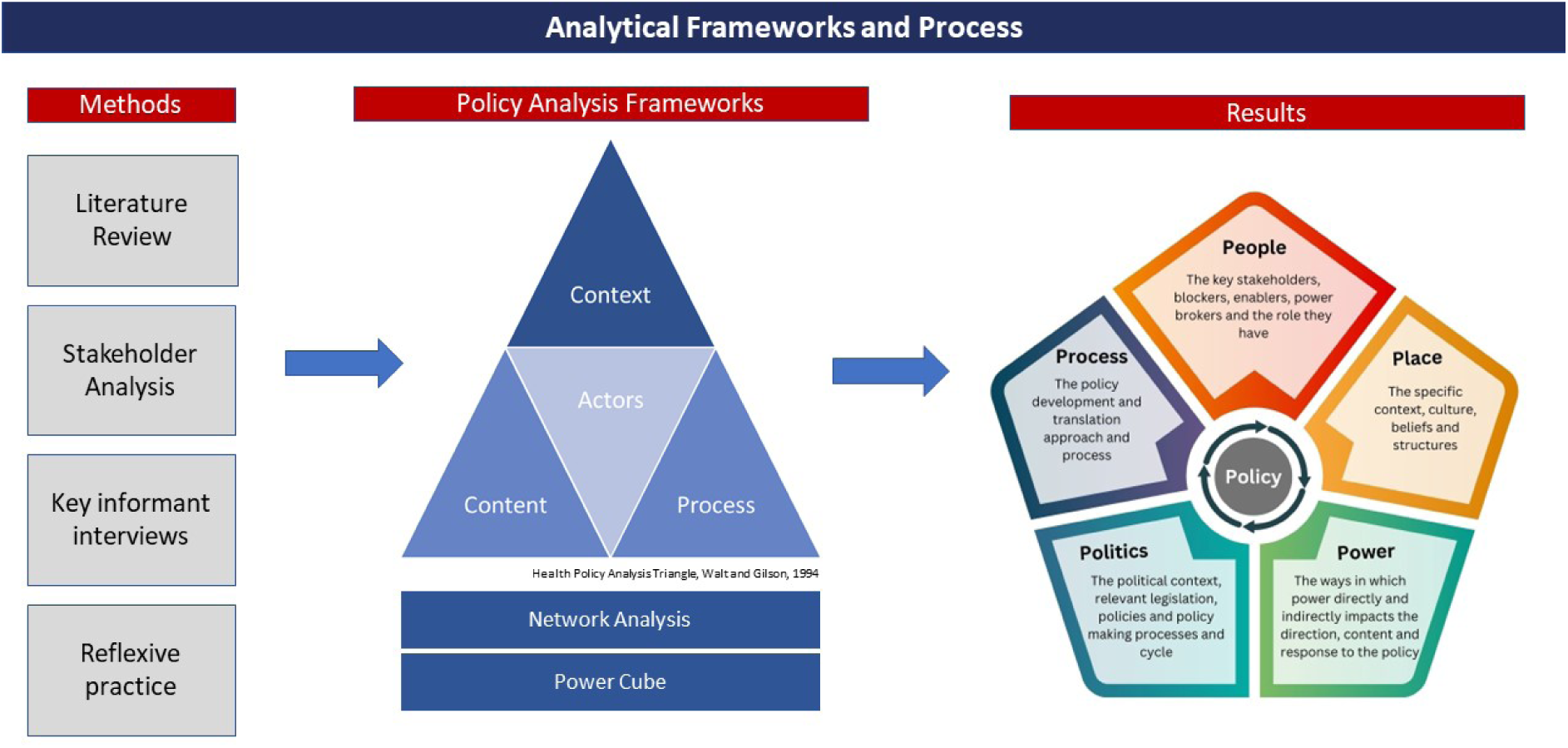
Analytical frameworks and process informing data analysis and interpretation.

### Ethics Statement

Ethical approval for this study was obtained from the Liverpool School of Tropical Medicine (#22-056) and the University of Liberia, Atlantic Centre for Research and Evaluation Institutional Review Board (#23-01-356). Prior to the interview, participant’s informed consent was obtained according to the protocol submitted for ethical approval prior to interviews.

## Results

The results are organised into five domains: people, process, place, politics and power (see Figure 3.) The dynamic relationship within and between the domains is critical to understanding the policy development and implementation process in the context of the integrated NTD policy in Liberia.

**Fig 3.**
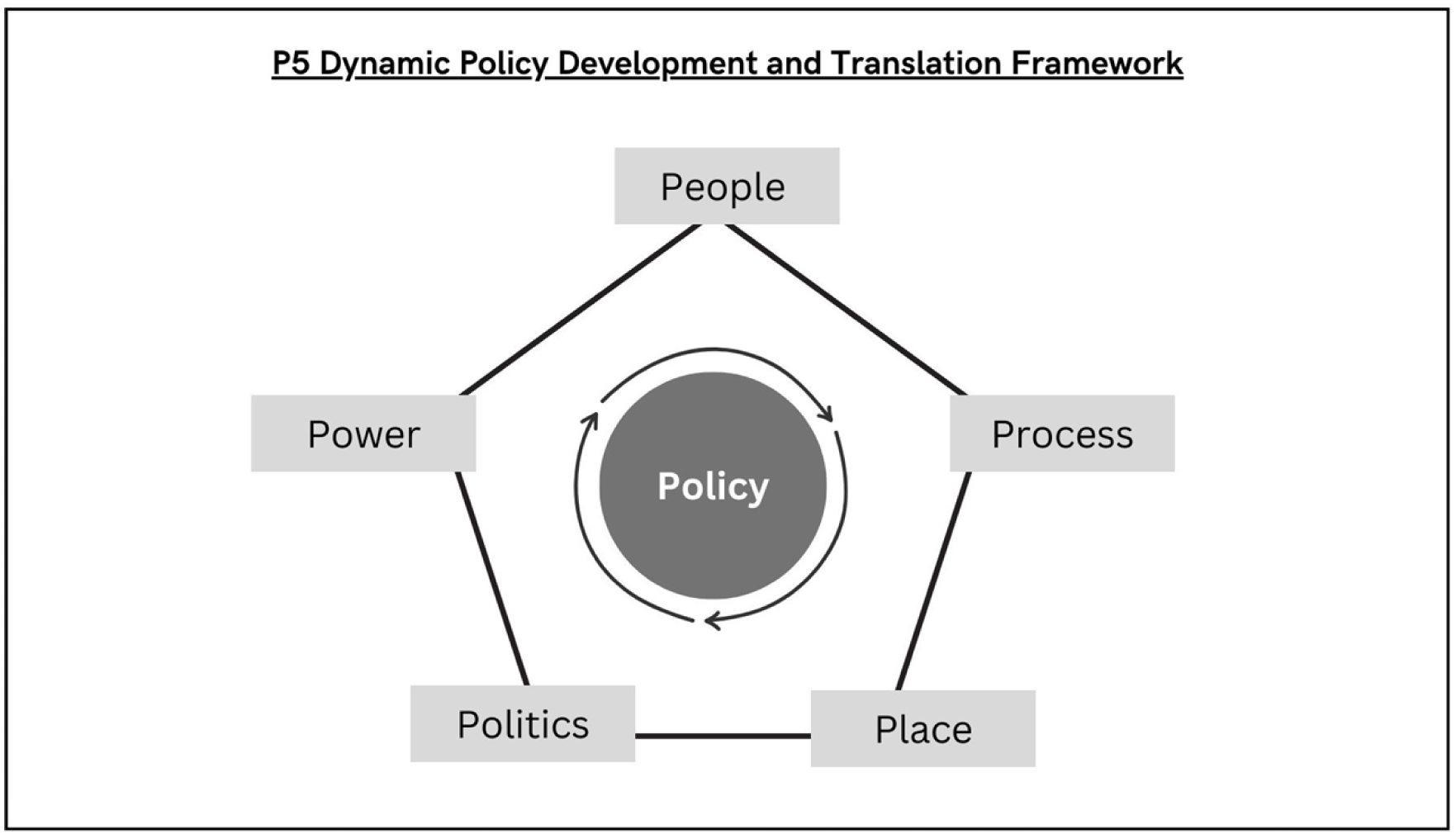
P5 Dynamic Policy Development and Translation Framework – Adapted from the Policy Triangle Walt et al 1994.

### People – The key stakeholders, blockers, enablers, power brokers

#### The impact of Ministry of Health leadership and ownership of the policy process

Key informants agreed that the ownership of this integrated approach to NTD case management belonged to the leadership of the NTD programme and senior policy makers within the Ministry of Health. Many stakeholders emphasised the importance of the leadership’s role and approach, with NGOs and donors particularly stressing the pivotal role of the NTD programme leadership in fostering political will, trust, and commitment to integration and encouraging collaboration between partners.

Moreover, visible support from senior ministers within the Ministry of Health at critical points in the development of the Policy, reinforced the Programme’s leadership and underscored the importance of stakeholder participation. This high-level backing also promoted integration across various Ministry of Health departments.

> “There are a few people at the government level…that are like that. So [the NTD Programme Director in Liberia] was one of them… they push the programme and the partners… I think it’s very good. I enjoy.. because I think as an organisation, you’re really there only to support, you’re not responsible, you’re not the implementer. You are there to support the process at country level…the country led model.” (International Key Informant −002)

Multiple stakeholders emphasised this point,

> “[The] leader of the NTD programme in Liberia from my perspective has been a very good leader, putting everybody together for meetings despite the tension of different partners to go by their [laughter] own, to fulfill their own agenda. But he has been very instrumental to try to put together all the partners.” (International Key Informant −005)

The importance of Ministry of Health ownership and leadership was also emphasised as a key factor in both the development and translation of the policy into practice by stakeholders at the national level.

> “It was driven by us. We know what we wanted to do. Unlike the Master Plan…this is something that we decided to do as a country…we knew where we wanted to emphasize; we knew our priorities, so even though, we adopted some things from the master plan… there were areas that we wanted to focus, and we did that.” (National Level Key Informant, Ministry of Health −003)

In an interview with a senior stakeholder within the Ministry of Health at the national level, it was argued that leadership was so critical that capacity strengthening should focus on empowering and equipping Programme Managers with management skills and training, beyond their clinical expertise to foster country ownership and leadership.

> “…the challenge now…it should be more like strengthening administrative skills. Creating a team of strong managers at all levels of the system, so they can know what to do.. [to]do it independently.” (National Level Key Informant, Ministry of Health −006)

#### Person-centred values and empathy driven decision making

A consensus among key informants suggests that shared values and a clear vision fostered an atmosphere of trust, collaboration and transparency. By emphasizing and modelling these values in critical relationships, sensitive topics like financial compensation, restructuring, financial commitments, and power dynamics could be tackled constructively. This values-driven approach promoted shared ownership, commitment, and consent for major shifts in power and programming models.

> “I always felt like the national team was very driven to ensure that…people were able to get the care they needed and weren’t being effectively neglected. That’s always the opinion I got from them.” (International Key Informant −005)

The importance of remaining focussed on the end goal of the policy rather than the barriers to getting there, was evident in many global and national-level interviews, it also resonated with stakeholders focused on the operationalisation and translation of the policy at lower levels of the health system. In one interview a stakeholder from the Ministry of Health described his feelings about the shift of leprosy from the TB Programme to NTDs. His description focused on what he saw as the route to a reinvigorated emphasis on leprosy in Liberia, which would serve the Liberian population better than the status quo, not his own position which had changed as a result of the policy.

> “Some people felt that they taking the programme from them… some people felt that it was good to do that, so that the patients can… have maximum benefits… when they saw the importance of it being like that … I think people felt good about it. And myself… I felt good about it…my satisfaction is the people suffering should be served properly … let the people have good services from their health workers, so if the people are benefiting, I am benefitting.” (National Level Key Informant, Ministry of Health −002)

Another key informant, whose role changed following the adoption of the integrated policy, was reflective about the values that must be kept in mind by anyone embarking on this sort of integration process, emphasizing collaboration and cooperation above the desire to maintain control and position.

> “So in order to improve the programme, if it needs to be restructured, we should accept it for the benefit of the health care delivery system because we cannot just keep it in our palm. It’s got to be expanded and to expand it we need knowledgeable people. People who have the knowledge to manage the programme or other areas of intervention, we need them… Yea, to help our people.” (National Level Key Informant, Ministry of Health −001)

#### The role of champions, capital and sacrifice

Personal and professional sacrifice to enable the development and implementation of the integrated approach is described by multiple stakeholders. The integration of NTD care within the Ministry of Health, the high level of ownership demonstrated by the NTD programme, and the greater harmonisation of NGO efforts and support was made possible by the commitment and sacrifice of key stakeholders who were significantly engaged in the policy development process and decision making. A key informant from an NGO described how their commitment to the transition from NGO led implementation of NTD care to Ministry of Health led implementation in Liberia had created significant challenges and problems with the senior leadership and the Board of his organisation, ultimately leading to the breakdown of the relationship between him and his supervisor and the loss of his job.

> “It was really difficult…I felt very bad about that because um… keeping explaining and you are not very well understood sometimes you have to find another way to make sure that you can continue to develop the passion and the vision that you have. And for those reasons, I have resigned and moved to another Organization which offers me the possibility to continue such moves.” (International Key Informant −004)

Other stakeholders talk about the challenge of the volume of work linked to a Programme with multiple diseases and translating this policy into practice within a sector where so much funding, measurement, and policy development is still reliant on a vertical disease-focused approach. They describe the huge volume of work that is created and how they try to manage the intersection that they find themselves at.

> “My reflection right now is … my role….comes along with lots of responsibilities and … you just have to…dedicate your services, because if you don’t dedicate your services, these things will not work. Because it’s an everyday activity, you don’t rest, yes you just don’t rest. And trust me there are times I come a long way and said well; at least this week I will rest, I have done X,Y, Z so let me rest and then tomorrow I will just have a phone call that needs my intervention… That requires either I move there or do some level of mobilization among the staff that I supervise to bring the situation under control. So, if you don’t put yourself in the state of readiness at all times that alone can compromise the implementation. I mean the integrated approach is not centered around one person; it’s the involvement of everyone.” (National Level Key Informant, Ministry of Health - 004)

Tied to ideas of personal and professional sacrifice is the role of social and political capital, especially evident among those deeply involved in developing and writing the integrated NTD policy. Interviews highlight how individuals, deeply committed to the policy’s vision, leveraged their social and political capital to gain support for integration and assuage concerns from those wary of the shift. One key informant who worked with an institution responsible for the case management of one specific disease, described the advocacy that they had to engage in, and the political capital they had to expend, in order to participate in the development of the integrated policy.

> “we always knew there was the need for it, it was just how to go about it was the trick… I remember the workshop, getting the integrated disease control strategy in place, I had to really push from my side to even be able to go. It was not something that was [considered] of importance.” (International Key Informant −005)

Similarly, advocacy and political capital were seen as critical in building internal buy-in within the Ministry of Health. When asked for the formula for success in this area a national NTD Programme stakeholder explained:

> “The first thing will be the Chief Medical Officer. He should be the first person to be engaged, from the Chief Medical Officer, through his office… then of course through that other people can come in, see how important, how good is the integration..” (National Level Key Informant, Ministry of Health −001)

Several participants mentioned the limitation that was a result of the lack of inclusion of people affected by NTDs, informal health providers and community members in the development of the policy, and its effective translation.

> One of the things that we didn’t do at the time, which I would do differently now, was that we didn’t get much involvement of persons affected by NTDs..if we had… we would have identified that their needs were beyond just the chemotherapies, were beyond just the management, the wound dressing, we would have understood strongly that they have psychosocial concerns, and we have wanted us to address those psychosocial concerns…I would get …their family members or caretakers. And I would get community stakeholders… I would get traditional healers, faith healers, and other community …have the plan to be largely influenced by their felt needs. (National Level Key Informant, Ministry of Health −007)

### Place - The specific context, culture, beliefs and structures

#### Health systems shocks creating momentum for change - the impact of Ebola

Liberia in 2015, was a country emerging from one of the most significant health crises of a generation, Ebola. Ebola had a catastrophic impact on Liberia, its population, its health system, and the trust that the population had in the health system’s ability to protect the health of its population. Ebola has highlighted not only the weaknesses of the existing health system but also demonstrated the feasibility and strength of integration and cooperation amongst multiple stakeholders alongside the critical importance of resilience within the health system.

> “Everyone came together from different sectors, different programmes. And I think that level of coordination and collaboration really opened up people’s eyes to the possibility of working together and the benefits of it, especially at a community level… And there may be a lot of benefits, as we saw with Ebola response.” (International Key Informant −001)

The discourse around the health system in Liberia shifted significantly following Ebola, the impact of a fragmented donor-reliant health system with little structural resilience had been exposed by the catastrophic impact on the lives of thousands of Liberians. National-level stakeholders discussed the importance of learning from the Ebola response, the shift in emphasis at the Ministry of Health and the increased understanding of how cross-departmental collaboration could work to achieve better health outcomes. However, some stakeholders believe the policy shift would still have happened without Ebola, but Ebola provided the opportunity to accelerate this shift. When asked if the policy change would have happened had the Ebola outbreak not occurred in Liberia, one respondent answered:

> “I am very sure it would have, it was because Ebola did not take away anything. It didn’t bring anything in the context of NTDs.” (National Level Key Informant, Ministry of Health −003)

#### Pre-existing health systems structures and policies - the irony of the opportunity of starting from zero

Something that multiple stakeholders mentioned, was the opportunity that was created by the absence or weakness of pre-existing vertical NTD case management programmes. Whilst seemingly counterintuitive, the absence of established strong and well-supported vertical programmes created an opportunity for integration with minimal resistance. When reflecting on the experience of the integration of leprosy, which did have a historical programme that predated the integrated policy and was combined with TB, key informants from within the Ministry of Health talked about how hard it would have been to have made integration happen if the process that had to be undertaken to integrate leprosy had also had to happen for the integration of the case management of other NTDs.

> “So I think part of the reason why we were able to pull the NTDs as an integrated programme was that most of them were not existing as a well established vertical programme. I just give you an example of how leprosy was, assuming all of the nine diseases or conditions were established vertical programmes… and we had to pull all of that together, the course that we pass through with leprosy, we would have had to pass through that with all of the diseases, that will be a huge, huge, huge challenge that we would have had to spend the next three years or four years trying to bring everybody on board. So, I think part of it was the fact that these programmes… were not full grown established programmes by themselves. So that was a blessing in disguise.” (National Level Key Informant, Ministry of Health −003)

#### The influence of traditional beliefs, culture and values

During data collection with county level informants an interesting observation that was made in response to the question, “why do you think integration was possible in Liberia?” was that it reflected deeply held cultural values, norms and practices of collaboration and working together to heal those members in the community that were unwell or vulnerable at that time.

> “Our cultures and values…. It makes you to teach your traditional behaviour, how to live within your community, how to treat people, how to [ensure] things are working together, especially in the community.” (County Level Key Informant, Ministry of Health-003-M)

#### Addressing the pressures on health care workers and policymakers in the context of a fragile state

Within the context of a fragile state, there are particular pressures on health workers that are often ignored or seen as being outside of the remit of a policy such as this. One of the areas that was most often identified as a critical area of the policy, was the discussion around, and inclusion of strategies to address, the challenge of unreliable and reduced salary payments.

> “I go out, I am one of the national trainers. I also received some benefits from the training, so for this….I say….It’s a plus for me as I was saying; if I am sent out and my family suffering behind me, I may not be satisfied there, but when the satisfaction is there, with the little I am getting, it will support my family also. Children going to school, and my family eat, you know.” (National Level Key Informant, Ministry of Health −002)

### Power - The ways in which power directly and indirectly impacts the direction, content and response to the policy

#### Influence of partners on policy change and implementation

When asked about influences on the adoption of the integrated approach respondents referred to the influence of the World Health Organization (WHO) and partners on the thinking that drove the policy shift, whilst also emphasising the ownership of the decision by the Ministry.

> “…partners supported [local NGO office] and started building capacity as well, yea including the programme manager travel and saw the need, yea, it was through partners who supported the programme managers who went and saw …the possible impact on the programme. So, partners supported, including WHO, for us to know, to accept the need, and to know the importance of integration.” (National Level Key Informant, Ministry of Health −002)

The power of partners to enable or create barriers to the policy was acknowledged from the outset; key informants described the effort and time invested to bring partners on board and engage with them throughout the process. This was undertaken to ensure that integration was not a shock and to mitigate resistance to the policy shift. Partners had the opportunity to participate in the discussion throughout the policy development process. Despite this engagement, there are examples throughout the data of some partners being resistant to the shift towards integration, influenced by their own concerns and interests, particularly regarding the visibility and impact of their financial contributions. Some key informants explained that Partners sometimes questioned the reputational or organisational benefits of their investment, concerned that their contributions would not stand out and results be recognised distinctly as attributable to their contribution. Consequently, there was pressure on the programme to adapt its plans and operations to align with what these partners believe is best.

There were also examples of positive approaches to partnership, and those partners that also invested In the NTD Programme with a longer-term strategy:

> “And then partners who have been supporting this case management plan have been consistently involved year after year. And not just with funding support, but also with technical inputs…And by doing this monitoring, evaluation and learning, together, we have tried to improve or stay on track on what the Policy was supposed to achieve. So, the consistency, I would say, is really important across the board.” (International Key Informant −003)

#### The role of funding in enabling and sabotaging policy change

The impact of the funding approach of partners was also emphasised as a critical factor in the translation of policy into implementation. With a high level of appreciation for partners and funding that was able to be responsive to the complexities that arose in the implementation and the need for adjustments in budgets and activities. It is also apparent the damage that was done to the implementation of the approach and trust that had been built by the decision development and approaches of some partners.

> “A partner’s funding changed in mid-year with activities already planned and budgeted for and agreed upon. And I think, sometimes partners are not mindful… they think of numbers and forget the actual people that are being affected. So, suppose you cut funding from a programme. In that case, those community health workers that are being transported to find cases may not be able to do that, which means there are more and more cases that won’t be found, increasing risk of disability for those patients, there are health workers that wouldn’t get refresher training that might go one, two years without seeing an NTD case. And will have to go back to square one because they will no longer remember how to clinically diagnose BU accurately and all those types of things.” (International Key Informant-001)

An additional aspect of funding related to its role ensuring the translation of policy into implementation. As part of the policy development process, existing partners were asked to share their funding ceilings and funding restrictions with all stakeholders. This was not to limit the scope of the policy development but meant that the momentum and buy-in that had been built during the policy development process were able to be leveraged and turned into implementation support. This did have limitations as the available funding was not sufficient to support the implementation of the entire strategy, but it did provide a bridge between the policy and actual implementation.

> “To have money available…I noticed that the moment these plans are launched, the programmes are still in the momentum, you’re still excited, the enthusiasm is still there, all the different partners or other programmes that were invited to the table there, they’re still keen on NTDs. When that programme is launched, and you wait three, six months, before you can have some small, not even adequate funding, some small funding available, you tend to lose the momentum…. NTDs is not all that popular already, so when you get people at the table who are like, okay, I’m interested, I’m listening, you have their attention, you want to go on there now.” (International Key Informant −001)

#### Negotiating the fear of redundancy, loss of control and the legacy of colonialism

At the foundation of many of the challenges and resistance that stakeholders perceived about integration, was not simply a power shift but also a fear of redundancy. When reflecting on this fear and the impact that it had, one stakeholder explained:

> “There were times that we met in the corridor and when we exchange greetings, they will say just go that way because you want to take away my leprosy. [laughter] You don’t speak to me again.” (National Level Key Informant, Ministry of Health −004)

Another international key informant reflected on the tension for NGOs of moving beyond a vertical disease focus:

> “If you look at leprosy, it’s the same people all the time, you know, still, like from 20 years ago, really to be honest, that’s the same people … that went from national positions to global positions… I think that the world is still too closed at the higher level, and then at the organisational level, they are also still doing what they have been doing, even the names… that refer to leprosy…so the whole organisations are, like, ingrained, it’s ingrained in their existence.” (International Key Informant −002)

The fear of redundancy and insecurity relates closely to the sense of a loss of power and control. Key informants describe multiple ways that this fear manifests, amplified by a lack of understanding or appreciation of the complexity of the process of integration, the distinction between co-implementation of activities at the field level and systemic integration and an impatience with the time that is required to integrate systemically into the health system and to demonstrate disease-specific results that can to some extent be attributed to this integration.

> “So I think it can be demotivating, I think sometimes partners, so to speak, bring the programme to their knees to beg. And it’s unfair, because most of the work, all of the work, has been done by the programme and the people in the field and… … as if you’re on trial, and you’re being questioned and questioned, are you sure this is happening… and the interrogation and I can tell partners for sure… if there was a case where the government will ever say we have money there, we’re just not even listening to partners, like a lot of partners will be dismissed because they wouldn’t be needed. They’re at the mercy, so that’s why they’re there, but it is demotivating to the programme because they see the impact [from what they are doing] and are excited about what’s happening. But then partners are questioning like, I don’t see anything like what’s happening. And it’s almost like a blame game at some point. And so it’s not encouraging at all.” (International Key Informant −001)

Power also manifests in the responses of participants in relation to priorities and agenda setting; several key informants also discussed the impact of external stakeholders, including global policymakers, on influencing priorities and agenda setting. Throughout the data, stakeholders from NGOs and donors talk about the challenge of a shift in organisational narratives that are so interlinked with the identity of many funders of the case management of NTDs and the ways that these organisational identities, often tied to specific diseases, challenge the understanding and capacity they have to communicate an approach, such as the integrated approach in Liberia. This challenge has led to a perception that the agenda and priorities of some NGOs and donors are not aligned with the direction of the Ministry.

> “I think, as NGOs, then you need to say, okay, my mandate is to support the national effort. You know, it’s not that I am not ending leprosy but I’m supporting the integration of these diseases. And with that, I’m supporting the process of ending four or five diseases instead of only one. So, your mandate needs to change. And that is, I think that that is difficult at the moment because the disease-specific organisations, they’re still pushing their own thing.” (International Key Informant −002)

### Politics - The political context, relevant legislation, policies and policy development processes and cycle

#### Global policies and their impact on agenda-setting and policy development

Key informants make multiple references to the influence of global and regional policies on the decision to move to an integrated approach to NTD care. Key informants describe both the positive influence that global policies, such as universal health coverage, IPCHS and the WHO NTD road map had, as well as the challenges that were caused by the apparent contradictory nature of some of the policies, which rather than supporting a shift to a health systems based and integrated approach instead supported either explicitly or implicitly an approach that maintained the status quo where vertical or parallel implementation of NTD programmes was maintained, and focus on specific diseases were dominant.

> “The integration at the country level is good. But at the international partner level, even at WHO it is still not there, I enter WHO office I see PCT NTD, I saw CM NTD, I don’t see integration happening… but they’re asking us to do integration. If you don’t integrate the human resources at that level, how do you expect that the resources, in terms of the financial resources are going to be integrated? So, if you are there at the top, there’s a challenge integrating, but we want to do it at the bottom. So, what are we doing to see which one is feasible at the moment. So just saying this to say that yeah, there will be always worry, especially when systems have power, unfortunately when they are parallel to bring them together is a whole lot of challenge.” (National Level Key Informant, Ministry of Health-003)

#### History and cultural ties to the status quo

One of the most contentious themes was the integration of leprosy within the NTD programme under the integrated NTD policy. There are multiple facets to this specific issue. However one of the key arguments made by stakeholders from within the TB and leprosy programme when asked for their reflections on the policy was that the integration of leprosy within NTDs violated legal statute which established the leprosy programme in Liberia and its subsequent partnership with TB in law and that due process had not been followed to address the need to change this law to reflect the policy change that was being pursued. Very few key informants raised this specific political and legal barrier, but it was a significant line of argument among some the TB/leprosy programme:

> “Okay, but he knows very well that the technical team or, the knowledge when it came to leprosy were sitting right at the National Leprosy and TB control programme. He knows the policy, he knows the Act that created the National Leprosy and TB programme… we will let them know that we…. want the Act amended so that it can take the leprosy programme and move it under the NTD programme” (National Level Key Informant, Ministry of Health −005)

#### The impact of the political context

The political context in Liberia at the time of the development of the policy and throughout its implementation has changed significantly. Elections in 2018 marked the change of administration in Liberia and significant shifts in the leadership and human resources within the Ministry of Health. When reflecting on the initial policy development process several stakeholders describe the positive impact that senior political leadership from within the Ministry of Health had on building buy-in and commitment to the policy shift. Whilst there was some continuity in the leadership at the Ministry of Health, there was also significant shifts in policies, specifically there was a policy referred to as the ‘harmonization policy,’ which was implemented following the election in 2018 with the reported intention of harmonizing government worker salaries, but which is also mentioned by several respondents as leading to an abrupt and dramatic reduction in salaries (29, 30), and having one of the most significant and negative factors in the translation of the integrated policy into practice. The political context is also described as a challenge for the communication of the priority of NTDs within the political system, key informants describe the need for continued and greater advocacy to facilitate and maintain the progress towards integration that has been made.

### Process - The policy development and translation approach and process

#### Rationale for the transition to an integrated approach to NTD case management

Key informants emphasised multiple factors that led to the shift in policy. The increasing profile of NTDs following the London Declaration(31), the sustainable development goals and universal health coverage(8), and emerging frameworks for health systems, such as the IPCHS (10), created a general policy direction that supported this shift from vertical service provision to an integrated model of delivery. There were also some important interactions between the National NTD Programme and influential stakeholders, such as WHO, immediately preceding the Ebola outbreak, where integration was recommended as a strategy that should be considered by the NTD Programme in relation to NTD case management, specifically leprosy and Buruli ulcer.

#### Participatory processes to build stakeholder buy-in, and mitigate unforeseen barriers

One of the elements of the policy development and implementation process in Liberia that was cited most often by stakeholders from national and international NGOs and other donors, was the participatory approach that was used in the policy development process.

> “Yes, I think we were together, and we developed also together, we, from the field perspective….the programme itself, we were together for the development of this first integrated plan, which was very, very important. And so this is not like a… me as an individual, it was like we were in pool of people putting together our idea to make it happen. I remember, this plan has been developed we had revised it several times in such a way that it meets the err..err.. agreement of all the people around the table before it goes” (County Level Key Informant, Ministry of Health-004)

Whilst the policy’s ownership lay firmly within the Ministry of Health’s NTD Programme and the senior ministers to whom the NTD programme was accountable, extensive consultation with and participation of other stakeholders from within and outside the Ministry of Health was not only encouraged but proactively sought.

> “The Ministry of Health really provided that sort of convening role between all the partners who wanted to work together and as we built this policy, it was apparent that it would require a lot of effort from everybody involved, especially the NTD programme. (International Key Informant −003)

Multiple stakeholders talked about the process’s impact on their ability to communicate the policy’s content and intention throughout their organizations. Whilst this did not mitigate all of the resistance to integration, it did provide tools needed to reassure and counteract fears and uncertainties among many stakeholders.

#### Leveraging a whole system-based approach

Respondents were asked, if the process was to be undertaken again, who should be included; the responses supported a highly participatory approach to policy development to enable the effective translation and implementation of the policy throughout the county, district and community level:

> “We have a county health board. So, we will invite the superintendent. You know, the county health board is chaired by the superintendent and secretary is the CHO. So, if we have that policy, I alone will not sit down and make the policy. We have to invite the county health board…. We invite the district health officers, the district surveillance officers and the head of the OICs to be in that policy development…so that we can develop one policy and we go by it.” (County Level Key Informant, Ministry of Health-001)

Another county level stakeholder added:

> “Cases are not just seen like just in the community, but within the same community, they have other structures there, that cases run to like the traditional herbalist. The traditional herbalist, they need to be informed about cases and even those religious people. We have some areas where the people have prayer people, praying for people to those areas, you see NTDs cases going there… we need the involvement of stakeholders in order to get the programme to be effective.” (County Level Key Informant, Ministry of Health-002)

#### Creating space for the complexity and process of integration through time

Finally, but of critical importance to the majority of informants was the need for an understanding amongst policymakers, partners and donors, as well as various health systems actors, that integration is a process that will take time, it does not end with the development of an integrated plan or strategy. Related to this, is the need for policymakers to think beyond disease specific targets to actually allow and enable the measurement towards integration and sustainability and to celebrate this as a real achievement in and of itself.

> “What are the targets country wise, not the national, the international level, like I listened to the guys for the WHO say, by 2030, a number of countries, have integrated plan… that is not what I am talking about. I am talking about impact, impact related, measurable indicators that can cut across all of the diseases. In my mind when you start doing that, then you know, you’re getting closer… to the way you’re paving for, for integration and being able to say we have done it and we can measure this.” (National Level Key Informant, Ministry of Health −003)

## Discussion

To develop and implement Liberia’s integrated NTD policy effectively, five critical domains were addressed throughout the development and policy translation process (Fig 4); the relationships between these domains are dynamic; they should not be viewed as separate silos of activity but as interrelated areas that vary in influence and importance depending on the context and time.

**Fig. 4.**
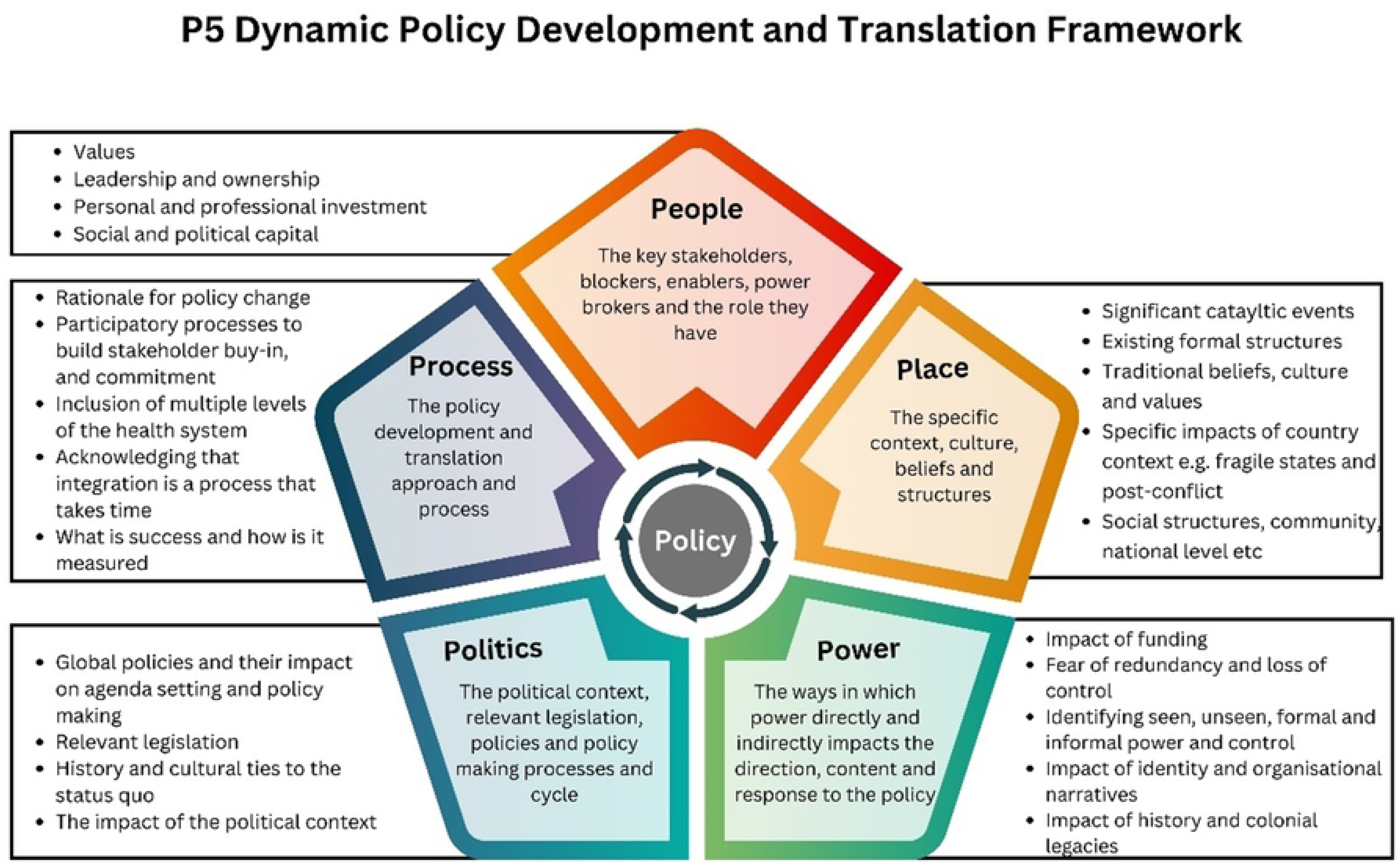
P5 Dynamic Policy Development and Translation Framework.

### People

This domain refers to individuals and key stakeholders related to the policy process. The leadership’s shared vision and values, the Ministry of Health’s ownership, the personal and professional commitment of critical figures, and their ability to use social and political capital to navigate challenges and power dynamics were all vital to the successful policy shift toward an integrated case management model in Liberia. The recognition and involvement of key stakeholders by the leadership and champions of the policy was also critical. The findings from the data in this domain support earlier results from the participatory stakeholder analyses: firstly, the importance of recognising that a person-centred approach to integration and broader health system strengthening requires the application of multiple lenses when considering key stakeholders. Secondly, the importance of engaging stakeholders at the subnational and local levels as those stakeholders are critical to the successful implementation of integrated programmes. Finally, the importance of employing effective and appropriate methods and engagement models based on stakeholders’ needs and positions. These methods and models are also related to the ‘Process’ domain and can be considered in three broad strategies: 1. informing and engaging; 2. consulting and inviting; and/or 3. relationship building and influencing. The importance of conducting a stakeholder analysis, utilising and then updating it throughout the policy development and implementation process is critical; this supports the literature on the importance of considering ‘actors’ in policy analysis (11, 13–15, 32), on stakeholder analysis (16, 33) and the practical emphasis that is put on stakeholder engagement by multiple policy-development and influencing institutions. (33)

### Place

Place refers to the environment where the policy was both developed and implemented; factors in this domain to consider include the specific cultural context, societal beliefs, and existing socio-economic structures within Liberia, which significantly influenced the policy’s development and implementation. Factors such as the impact of the Ebola outbreak and the exposure of the weaknesses in the existing structures that required integration played pivotal roles (34–36). Furthermore, the influence of traditional beliefs and cultural values related to conceptions of health and the policymakers’ willingness to address the unique strains on healthcare workers in a fragile state context were also significant. This domain should also consider the specific socio-cultural, demographic, or physical places that are being targeted through the policy change and the place where the policy will be implemented, such as communities and health facilities. The consideration of ‘Place’ in policy analysis has been recognised in multiple research areas, from health policy analysis frameworks (11, 13, 37) to the emerging literature on culture and health and the intersection of these concepts (38, 39).

### Power

The dynamics of power unmistakably shaped the policy development process and its translation. This includes the influence of partners on policy change, both positively and negatively, the role of funding availability in the translation of policy into practice, the fear and uncertainty linked with policy change towards integration and greater government ownership, the loss of control felt by some stakeholders, and the effects of entrenched colonial attitudes and disease-specific organisational identities.

The findings from this study support the literature on policy development and the political economy that has been emerging since the early 1990s, especially building on the findings that power and process are critical to policy and policy change (40, 41). The other related body of literature that is significantly supported by these results relates to the importance and role of power analysis within the context of both health policy and development policy (26, 42–48).

Within both global health and development studies, there is a growing appreciation of the impact of colonisation and related power dynamics on the conceptualisation of global health programmes, policy-development and implementation processes in low- and middle-income countries (26, 43, 46, 49–52). One of the most striking facets of the policy-development process was the extent to which power was transferred away from traditional colonial models of healthcare provision through the expenditure of social and political capital by critical stakeholders, and the techniques and strategies that were employed to enable this transfer of power, priority and agenda setting. However, the data also describes the ongoing legacy of the prevailing power dynamic and how these dynamics continue to create barriers to national ownership and agenda-setting for NTDs.

### Politics

This domain describes the political landscape, relevant legislation, policy-development processes and cycles, and how a specific policy fits within the broader national policy environment. It also considers the global and regional policy context and its influence on national agenda-setting and policy-development. Factors such as election cycles, inadequate budgetary allocations and legislative priorities are also political considerations. The importance of these political processes in many ways reflects the conception of health as an innately political concept. (41) This concept is explored by Bambra et al. (40), who describe the political nature of the concept of health.

> “Health is political because, like any other resource or commodity under a neo-liberal economic system, some social groups have more of it than others. Health is political because its social determinants are amenable to political interventions and are thereby dependent on political action (or more usually, inaction). Health is political because the right to ‘a standard of living adequate for health and wellbeing’ (United Nations, 1948) is, or should be, an aspect of citizenship and a human right. Ultimately, health is political because power is exercised over it as part of a wider economic, social and political system” (41)

This is especially pertinent in understanding the complex policy process that was undertaken in Liberia to integrate NTDs within the health system. NTDs, by definition, impact the most neglected populations, are neglected in global health funding, research and policy and are deeply linked to issues of inequity, so to overcome these systemic and structural challenges and inequities, understanding and addressing power is critical.

### Process

This domain explores the policy development and translation process itself. The data supports the literature that emphasises the value of understanding the reasons for policy change, the importance of participatory processes for building stakeholder engagement, trust and commitment, the necessity of including diverse perspectives, and the inclusion of actors at multiple levels of the health system as well as those with lived experience (53, 54). It also recognises that policy development and implementation are ongoing rather than one-off events and that success may look different at various stages of the policy process and should be measured in a way that reflects that. A significant body of literature exists in the field of development studies and research that emphasises the critical role of participation in the implementation of development interventions and policy development. However, there are few empirical studies of the role of participatory processes in the development and translation of complex global health policy (54–57).

### Controversies around the definition and application of integration for health systems strengthening

The 1978 Alma-Ata Declaration (58) marked a pivotal moment in 20th-century public health, championing primary health care as the means to achieve ‘Health for All.’ Implicitly and explicitly, it stressed the importance of integration within the health system. However, despite recognition of the need for integration dating back to Alma-Ata in 1978 and its prevalence in public health discourse, no universally accepted definition of integration exists. This lack of consensus has confused its application and the lessons learnt from integration efforts. The term “integration” has spawned over 176 different definitions, shaped by various stakeholders’ perspectives, including clients, providers, policymakers, funders, and evaluators and was sighted by policymakers in Liberia as a barrier to the development and implementation of the policy (21, 23).

Within the World Health Organization (WHO), there are multiple definitions of integration, all describing phenomena that bring together different elements or functions of health systems. A common thread across these definitions is their focus on placing the people affected or the broader population’s needs at the centre, emphasising collaboration and normative integration. Normative integration involves aligning and harmonising various system elements according to shared values, goals, principles, and standards (21, 59, 60).

The debate surrounding integration primarily revolves around integrating disease-specific interventions into health systems. It has been framed as a binary struggle between integrated (horizontal) and non-integrated (vertical) programmes, with proponents on both sides. (61) The primary rationale for moving towards an integrated approach stems from criticisms of vertical programmes, suggesting that their gains are inequitable and unsustainable without broader health systems strengthening or may come at the expense of the broader health system. (62) This has led to the emergence of a dual-objective implementation model that combines disease-specific vertical programmes with health systems strengthening (horizontal), often termed a “diagonal approach” (63).

### Lessons learnt and broader applications

An effective policy development and implementation process for integrating neglected tropical diseases (NTDs) into the health system is crucial for achieving global health goals related to NTDs and universal health coverage (1, 2) and the WHA resolution on people-centred integrated health services (3). There is minimal evidence in NTDs on how to do this effectively. (4–6) The experience of Liberia in this area provides critical insights that can significantly contribute to bridging the evidence gap in this domain and can inform a framework applicable to policy planning and implementation in various contexts; the purpose of this study was to explore this experience and situate the learnings within the broader literature.

The findings from this study can have multiple impacts to enhance policy-development processes for complex integrated policies and for increasing the likelihood of policies translating into practice. Specifically in relation to NTDs; there are essential indicators within the NTD roadmap in relation to the integration of Skin NTDs, including the target of 40 countries adopting and implementing integrated skin NTD strategies based on local endemicity by 2030 (64). Understanding the learning from the process in Liberia could provide valuable guidance for moving further towards this target amongst many others related to reorienting the health system to mainstream NTDs.

These learnings are not only restricted to NTDs, but they also provide important reflections for countries seeking to achieve Universal Health Coverage and move beyond parallel and vertical approaches to do this, especially in fragile states and health systems that are deeply dependent on donor funding and financial aid. The findings also have implications for thinking around capacity strengthening, resource mobilisation and measurement moving forward, providing evidence for the position that capacity strengthening and policy-development processes need to think far beyond disease-specific clinical expertise, but rather need to consider management capacity, flexibility in financing models, political advocacy skills and processes.

In the future, the findings need to be tested to ascertain transferability to other contexts. There also needs to be further research done on the meaningful inclusion of community members and people with lived experience in the policy development and translation process. There is also scope for a deeper application of an intersectional lens to policy analysis, using tools such as the Intersectionality-Based Policy Analysis (IBPA) Framework, which would significantly enhance this study.

### Strengths and limitations of the study

The strength of this study is how it has been able to examine one policy in depth from multiple perspectives. The insider status of the lead author and the collaboration within the REDRESS research project facilitated a high level of access to critical stakeholders representing multiple perspectives, all with a high level of engagement in the policy process that was the subject of this study. This study does have limitations. However, the focus on a single country means that the results are highly contextual. The insider status of the lead author, her previous and current work with the NTD Programme on behalf of an NGO, and her perspectives, could lead to information or selection bias given her positionality and be a limitation to the study and the ability of critical informants to participate with complete openness in the study. There is also a limitation that the key informants interviewed only include the perspectives of international, national and county-level stakeholders; this is reflective of this specific process of policy development which focused primarily on engagement at these levels. However, this in itself is a limitation of the study as no people affected by NTDs were included in the study. There is also a potential risk of bias with the inclusion of some key informants as co-authors, to mitigate this these co-authors were interviewed according to the ethics protocol and were not involved in the coding of their own transcripts.

## Conclusion

Policy processes in low-resource settings seeking to achieve the integration of vertical or parallel disease control programmes into health systems need to embrace complexity and make the essential shift from clinical-centric policies to holistic, multi-stakeholder strategies to policy development and translation in global health; no single NTD department or group of stakeholders can hold the health and wellbeing of people at risk or living with NTDs in the “palm of their hand”.

## Data Availability

Data are available on reasonable request

## Acknowledgements

We would like to thank all the participants who took the time to engage with us to be a part of this study and their accounts of their personal experiences. At the national and county levels, we thank programme managers, their supervisors and county-level health workers who served as key informants. We also appreciate key international partners, for the ongoing support and participation in the research. The REDRESS Programme (NIHR2001129) is acknowledged for the technical and other support provided.

## Notes

### Competing Interest Statement

The authors have declared no competing interest.

### Funding Statement

This study was funded by the National Institute of Health Research through the REDRESS Implementation Research Project (NIHR2001129). The authors (AW, ST, LD, ER and KK) received funding as part of this award.

### Author Declarations

Ethical approval was given by the Liverpool School of Tropical Medicine Research and Ethics Committee and the University of Liberia Institutional Review Board

